# Clinical factors associated with urinary albumin-to-creatinine ratio among pregnant women with preeclampsia

**DOI:** 10.64898/2025.12.31.25343297

**Authors:** Theresia L. Mboya, Monica L. Chiduo, Boniphace M. Sylvester, Kelvin M. Leshabari

## Abstract

**Objective:** To analyse clinical factors associated with urinary albumin-to-creatinine ratio among women with preeclampsia.

**Design & Methods:** A prospective cohort analysis was conceived at Amana referral regional hospital. Women with preeclampsia (exposed group) were compared with women without evidence of preeclampsia (control group) from any time after 20^th^ week of pregnancy to at most 24-hours post-delivery. A simple random sampling with 1:1 matching was used to get study participants. Each participant was admitted for a duration of at least 24-hours at baseline for estimation of urinary-based albumin-creatinine ratio via 24-urine collection. Clinical Report Form was the main tool for data collection. A multivariable linear model was set for final analysis after appropriate linear model assumption validation.

**Results:** We successfully recruited and prospectively analysed a total of 55440 women-hours of follow-up between January to June 2025. Systolic blood pressure (A.O.R.: 1.01, 95% C.I.: 1.00 – 1.77), diastolic blood pressure (A.O.R.: 1.12, 95% C.I.: 1.00 – 1.97), Caesarean delivery (A.O.R.: 1.19, 95% C.I.: 1.02 – 1.45), neonatal birth weight (A.O.R.: 2.0, 95% C.I.: 1.2 – 2.9) as well as newborn’s 5^th^ minute Apgar score (A.O.R.: 1.07, 95% C.I.: 1.0 – 1.33) were factors significantly associated with maternal urinary-based albumin-to-creatinine ratio in this study population.

**Conclusion:** Diastolic blood pressure had a higher risk than systolic blood pressure to predict significant proteinuria. Newborn’s birth weight as well as 5^th^ minute Apgar score were immediate outcomes associated with maternal significant proteinuria.

**Recommendations:** Fetal maternal calculators for predicting/prognosticating significant proteinuria among pregnant women with proteinuric preeclampsia are warranted.

## Introduction

Preeclampsia along with other hypertensive disorders of pregnancy remain among significant contributors of maternal deaths worldwide (1, 2). It refers to a new-onset hypertension with-(out) *significant proteinuria* and/or end-organ damage(s) associated with uteroplacental abnormalities (3). The pathology is characterized by the abnormal vascular response after placentation that leads to functional changes such as increased systemic vascular resistance, enhanced platelet aggregation, activation of the coagulation system, and endothelial cell dysfunctions (3). At present, there are several challenges associated with the syndrome of preeclampsia. In fact, even the standard definition of preeclampsia is obscured by lack of objectively based domain(s) from obstetrics viewpoint. For instance, even though preeclampsia was customarily referring to *antepartum* events, there is wider acceptance among practicing obstetricians, for the new observation of the syndrome to occur even after delivery (the so termed “*postpartum preeclampsia*”). Moreover, globally - the spectrum of the syndrome severity – including what to be included/excluded, in the clinical definition of *severe preeclampsia,* is a subject of much debates among obstetricians in both sides of the Atlantic to date (4, 5). This article focuses on factors associated with *significant proteinuria* in the clinical diagnostic pathways of preeclampsia.

There is potential evidence to suggest that pregnant women with preeclampsia should be wary of the occurrence of adverse pregnancy outcomes when presented with massive proteinuria (6, 7). However, significance of proteinuria in pregnancies seen in preeclampsia is an unfinished agenda among both obstetricians and diagnostic medical scientists (8-10). First and foremost, even though the gold standard method to detect *significant proteinuria* in pregnancy is a 24-hours urine test, the test takes time (whole day) to collect, and is therefore almost always not available to guide clinical decision upon first evaluation, that happens as an obstetric emergency/urgency (8). Second, other available tests – like urine dipstick tests, measure concentration of protein in urine but are also susceptible to fluctuations in the water contents of the urine (8). Third and maybe the most adverse is the fact that even with the gold standard test for urine protein estimations, is the fact that threshold for pathologic states of urine protein tend to be different across pregnancy duration, further complicating against a single value for usage in obstetrics. For instance, there are still debate on the clinical utility of preterm vs term preeclampsia in pregnant women, and whether in each case, the threshold of *significant proteinuria* should be the same or different (9, 10). All these diagnostic challenges for detection of *significant proteinuria* in pregnancy call for alternative tests/methods.

Several factors have been reported to be associated with *significant proteinuria* in preeclampsia (11-13). For instance, a decade long, registry-based study from Finland suggested that advanced maternal age at first delivery to be associated as a risk for *proteinuric* preeclampsia (11). Specifically, they reported that those women over the age of 35 years had a higher risk compared to women under-35 years (9.4% vs 6.4%, p<0.001) of developing *proteinuric* preeclampsia (11). Besides, the same cohort (women >35 years) had higher rates (19.2%) of preterm (<37 weeks) deliveries (aOR: 1.39, 95% C.I.: 1.24 – 1.56) as well as Caesarean section (50%) (aOR: 2.02, 95% C.I.: 1.84-2.2) compared to younger women deliveries (11). However, the findings were derived out of national data of hospitals registry, and hence all deficits/biases/errors prominent in hospital data/statistics could not be ruled out. Moreover, the analysis included not only women identified with preeclampsia, but also those with eclampsia (11). Previous published reports showcased *biases* and *random errors* associated with inclusion of significantly different disease stages (in the Finnish case, *preeclampsia/severe preeclampsia* vs *eclampsia*) in evaluating efficacy and accuracy of disease processes and/or diagnostic tests (14-16). Thus, the need for fresh evidence from data-based, preferably prospective cohort design!

Conversely, earlier published studies reported utility of urine-based protein-to-creatinine ratio as well as albumin-to-creatinine ratio as alternative approach in the assessment of *significant proteinuria* in preeclamptic women (12). However, their estimates were based on findings that could not distinguish utility of the test on different variants (e.g. *preeclampsia* vs *severe preeclampsia*) and/or differential pretest probability of preeclampsia, that have been reported to significantly affect test performance of proteinuria in preeclampsia (17). Besides, in almost all of them, they were characterized by relatively smaller sample sizes, and hence reduced power to detect significant differences if present. Thus, we designed a prospective cohort observational analysis to account for factors associated with diagnostic test performance of urinary-based albumin-to-creatinine ratio for detection of *significant proteinuria* in the diagnosis of preeclampsia.

## Design & Methods

We exclusively targeted pregnant women with clinical features of preeclampsia (*exposed group*) versus expectant mothers without preeclampsia (*control group*) for factors (maternal, neonatal & hospital) associated with urine albumin-to-creatinine ratio in the diagnosis of preeclampsia. Preeclampsia was defined on clinical grounds to any woman with hypertension (SBP ≥ 140 mmHg and/or DBP ≥ 90 mmHg) having either *significant proteinuria* (urine protein level ≥ 0.3g/24 hours or urine albumin-to-creatinine ratio > 0.3) or clinical features consistent with end-organ damages (e.g. *renal* – serum creatinine > 90mmol/L; *liver* – elevated transaminases > 2 S.D. of upper limit with (out) right upper quadrant of epigastric pain; *neurological* – headache and/or persistent visual scotomata; *haematologic* – thrombocytopaenia of <150 x10^9^/µL) with-(out) uteroplacental dysfunction (i.e. abnormal umbilical artery wave form analysis and/or foetal growth restriction) commencing from at least 20^th^ week of gestation.

The study was a prospective-cohort, observational hospital-based analysis, conducted at a 600-bed capacity (including obstetrics & gynaecology) Amana regional referral hospital in Dar es Salaam, Tanzania. Dar es Salaam is the business capital of the united republic of Tanzania and currently estimated to be among the fastest growing cities not only in Africa but also globally, including its demographic transition process (18, 19). Follow-up of each woman in the study was from anytime from 20^th^ week of gestation to at most 24-hours post-delivery provided that inclusion criteria were met. It took place between January to June 2025. Gravid women were included if they had at least 20 weeks of intra-uterine gestation (measured from the 1^st^ day of the last normal menstrual period), attended at either antenatal care or admitted at Amana hospital’s antenatal/labour wards. All gravid women with any known/established chronic diseases prior to the index pregnancy, those with imminent eclampsia as well as those with *non-proteinuric gestational hypertension* were excluded. Non-exposed group members had to meet all eligibility criteria in exception to being free from pre-eclampsia.

A simple random sampling technique using table of random numbers with final sample matched in 1:1 was used to get the minimum sample size required for the study. Specifically, a minimum of 518 randomly obtained gravid women were found to have at least 84% power under a priori α-level of 5% assuming the estimated preeclampsia prevalence rate of 9.5% from a study done in similar settings just recently before (20). Moreover, allowing for an additional 10% of the minimum sample size to cater for (unit or item) *non-responses* or lost to follow-up, the final set minimum sample size was at least 570 pregnant women. Thus, a minimum of 235 pregnant women with preeclampsia were to be compared with 235 pregnant women free from preeclampsia in order to answer the research question. Matching was arrested at diagnosis level (i.e. preeclampsia vs no preeclampsia) and gestational age (+/-1 week) only by design. Matching was purposely done that way, in order to leave a room for all other significant antecedents to be gauged for their potential to serve as associated factors during analysis.

A Clinical Report Form (CRF) was the main tool of data collection. It consisted of four different sections, namely – maternal socio-demographics + previous/current obstetric status (section I); maternal physical examination and laboratory findings (section II); maternal pre-labour & labour status (section III) as well as maternal & neonatal postpartum status (section IV). Of special interest to this paper, is the fact that urine albumin level, urine creatinine level was used to compute for the outcome variable (i.e. urine *albumin-to-creatinine ratio*) that was coded as *a continuous ratio variable data*. Performance test characteristics on likelihood ratios, predictive values, sensitivity and specificity have been reported and published separately (21). Prior to actual data collection, the CRF was pilot tested at Amana’s emergency (*casualty*) unit and included test-retest reliability assessment as well as content and construct validity assessment of CRF items. Data collection was done by the PI herself as well as five research team members, composed of 1 house officer in obstetrics unit, 2 nurses (1 – antenatal ward and 1-labour ward) as well as 1 laboratory scientist. Research team members underwent a 2-days workshop on good clinical and research practice as well as a *drill test* for quick/emergency management of preeclampsia/imminent eclampsia at hospital settings.

Each participating woman had her date of birth (recorded as DD/MM/YYYY), gravidity, parity, age at first gestation, marital status, highest level of education reached, occupation, referring facility, gestational age at entry (in weeks), family and social history, drugs history as well as number of living children stata recorded immediately after acceptance to participate into the study. Besides, maternal weight (in kgs), height (in cms) were measured and their values stored until analysis time. Maternal sitting Blood Pressure (BP) was measured after 5 minutes of rest using manual Sphygmomanometer (*Riester*™, Germany) and stethoscope (*Littmann*™, USA) with cuff wrapped-up around the left upper arm, a bottom edge about 1 inch above the elbow in two occasions at least 5 minutes apart. Each systolic and diastolic BP was approximated to the nearest integers. The arithmetic average of the two measurements was recorded in the spreadsheet as the final sitting BP measurement. Those with blood pressure < 125/85 mmHg were considered *normotensives*, those with BP between 125/85 to 139/89 mmHg were considered *pre-hypertensives* and those with BP greater than or equal to 140 /90 mm Hg were considered *hypertensives*.

A special category for *severe preeclamptic* was considered for all women whose sitting BP ≥ 160/90 mmHg with either evidence of significant proteinuria and/or evidence of end-organ damage(s)/uteroplacental dysfunction(s). Other obstetric data included maternal fundal height at baseline, maternal fundal height at labour/near delivery time, obstetric USS (fetal viability, fetal lie, features of intrauterine gestation), duration of labour as well as drugs history. Date of birth was subsequently converted to actual calendar age at the time of analysis. Maternal calendar age, systolic-diastolic BP, calendar age at first gestation, gestational age at entry, gestational age at delivery, weight, height, urinary albumin, urinary creatinine levels were treated as a continuous interval variable. Gravidity, parity, number of living children, neonatal 1^st^ and 5^th^ Apgar scores were treated as ordinal variables. The rest of the variables (including their associated levels) were subsequently analysed as nominal variables with appropriate/clinically-relevant dummies where necessary.

Laboratory analysis that included 24-hours urine volume collected from a mid-stream urine via a sterile urine container (Abbott™, Abbott Laboratories, IL - USA). Each woman was instructed to begin urine collection at a specific recorded time, empty her bladder completely, and discard the first and last urine samples to avoid contamination. For the next 24-hours, each woman was followed-up during urine collection in a large volume sterile container. Each woman subsequently ended up the urine collection exercise just prior to or at exactly the same time the next day. Out of the total sample, at least 60 milliliters of urine were put in a small sterile urine container – well labeled with participant number (instead of name), date and time of collection then sent to the hospital laboratory for 24-urine for protein and UACR levels. Both the 24-hours urine protein levels as well as urine albumin-to-creatinine level analysis were performed using Abbott™ Affinion 2 analyser (Abbott™, Abbott Laboratories, IL-USA).

Data analysis was preceded by initial exploratory data analysis. Prior to data analysis, initial data were cleaned and checked for errors every day during the data collection exercise. The activity was performed by the Principal Investigator herself. It mainly involved checking missing data, and whether there were errors in collection/entry or storage. Exploratory data analysis formed the initial stages of main data analysis. It involved analyzing data for important trends/patterns as well as summarizing data accordingly. Data from questionnaires was entered and analyzed using SPSS software version 25.0. Continuous data were summarized using median and interquartile range while categorical data were summarized using counts and proportions. With the use of the 24-hour urine results as the gold standard, cut-off value of 0.3 mg/mmol in urine albumin-to-creatinine level was used for prediction of *significant proteinuria*.

Multivariable binary logistic regression analysis was fitted to account for the association between the albumin/creatinine ratio and a number of independent variables after appropriate validation of linear model assumptions. Specifically, the outcome variable in SPSS software was dichotomized (with a cut-off point of 0.3 mg/mmol) with 1 (≥0.3 mg/mmoL – signifying *significant proteinuria*) and 2 (<0.3 mg/mmol – control-*non proteinuric*). Before fitting the multivariable binary logistic regression model, each independent variable was assessed singly against the dependent variable (UACR) for *linearity, homoscedasticity, significant multicollinearity, significant autocorrelation, independence* as well as *normality* assumptions. Besides, a test of interactions (effect modifications) were also performed to known co-factors but yielded non-statistically significant findings. All univariate analysed variables were considered candidates for multivariable analysis if they attained a p-value threshold of ≥ 0.2. Receiver Operator Characteristic (ROC) curves was evaluated to determine an optimal albumin/creatinine ratio value that maximizes sensitivity and specificity in the identification of significant and severe proteinuria that was based on 24-hour urine collections. An alpha-level of 5% was used as a limit of type 1 error in findings.

Ethical clearance for this clinical research was sought from the IREC of Kairuki University. Permission to collect data at Amana regional referral hospital was requested from the office of the municipal director of Ilala as well as hospital director at Amana hospital. All women were approached with a written informed consent prior to inclusion into the study. Informed consent included the goal of the study, potential risks and benefits of being a study participant – e.g. *possibility for a false positive/negative lab results*, a clause that participants had a choice not to respond/answer/test any/all variable(s)/query(ies) without affecting the care being received at Amana hospital as well as a clear address to where to send their correspondence in case of any adverse events. Besides, all study participants who subsequently developed eclampsia were managed in accordance to the locally available hospital protocol at Amana regional referral hospital.

## Results

We successfully recruited and prospectively analysed a total of 55440 women-hours of follow-up between January to June 2025. They comprised of 288 women with preeclampsia (*exposed group*) versus 288 women without evidence of preeclampsia (*control group*) from at least 20^th^ week of gestation to at most 24-hours post-partum. Women in the preeclampsia group had a median gravidity and parity of 2 (IQR: 1-4) and 1 (IQR: 0-2) respectively. Likewise, the median gravidity and parity of women without preeclampsia (control group) were 2 (IQR: 1 – 4) and 1 (IQR: 0 – 3) respectively. Otherwise, their comparative baseline characteristics are shown in table 1 below:

**Table 1:** Baseline characteristics of followed-up pregnant women at Amana regional referral hospital (January 2025-June 2025).

| Variable name | Continuous |  | variables |  | Mann-Whitney U statistic (df) | p-value |
| --- | --- | --- | --- | --- | --- | --- |
|  | Preeclampsia Median | Group IQR | Control Median | Group IQR |  |  |
| Maternal age (in completed years) | 28 | 24 – 33 | 27 | 23 – 32 | 40112 (574) | .18 |
| Gestational age (baseline-in weeks) | 24 | 23 - 26 | 23 | 22 - 25 | 36895 (574) | .03 |
| Gestational age (at delivery – in weeks) | 38 | 36 - 38 | 38 | 38 - 39 | 41230 (574) | .41 |
| Maternal systolic BP ( mmHg) | 147 | 146– 147.6 | 111.5 | 105 - 124 | 82944 (574) | .000 |
| Maternal diastolic BP (mmHg) | 94.5 | 91.5 - 98 | 61 | 61 – 72 | 81210 (574) | .000 |
| Urine Albumin-to-Creatinine Ratio –UACR | 3.7 | 2.7 – 5.1 | 0.31 | 0.12– 0.55 | 79840 (574) | .000 |
| BMI (kgm<sup>-2</sup>) | 28.5 | 25.8 – 32.8 | 25.5 | 24.4– 26.8 | 75910 (574) | .000 |
| Neonatal birth weight (kgs) | 2.75 | 2.4 – 3.2 | 3 | 2.7 - 3.4 | 35640 (574) | .02 |
| Neonatal Apgar score (1<sup>st</sup> minute) | 8 | 8 - 8 | 8 | 8 – 8 | 41472 (574) | 1.00 |
| Neonatal Apgar score (5<sup>th</sup> minute) | 10 | 10 - 10 | 10 | 10 – 10 | 41472 (574) | 1.00 |
| Categorical |  |  | Variables |  |  |  |
|  | Preeclampsia Count | group (%) | Control Count | Group (%) | Chi-Square* (df) | p-value |
| **Marital status** |  |  |  |  |  |  |
| -Single | 34 | 11.8 | 30 | 10.4 | 1.27 (2) | .53 |
| -Married | 251 | 87.2 | 255 | 88.5 |  |  |
| -Divorced | 3 | 1.0 | 1 | 0.3 |  |  |
| **Gravidity** |  |  |  |  |  |  |
| -Primigravidae | 87 | 30.2 | 92 | 31.9 | 0.2 (1) | .65 |
| -Multigravidae | 201 | 69.8 | 196 | 68.1 |  |  |

**Note:** \*Pearson $\chi^2$ -test was used to test for independence under 5% level of significance

Besides, the maternal groups (preeclamptic vs control) were also compared to their immediate neonatal outcomes in order to assess for any significant differences. Table 2 below displays the summary statistics of the comparative statistics of maternal groups and immediate neonatal outcomes.

**Table 2:** Comparative statistics of followed-up pregnant women and their immediate outcomes at Amana regional referral hospital (January 2025 – June 2025).

| Neonatal outcome | Pre-eclamptic group |  | Control group |  | X<sup>2</sup>-test (df), p-value |
| --- | --- | --- | --- | --- | --- |
| **Mode of delivery** | **N=288** | **%** | **N=288** | **%** | **56.3 (2), 0.000\*** |
| SVD | 97 | 33.6 | 187 | 64.9 |  |
| CS | 189 | 65.6 | 100 | 34.7 |  |
| Assisted | 2 | 0.7 | 1 | 0.44 |  |
| **Sex of neonate** |  |  |  |  | 0.11 (1), 0.74 |
| Female | 135 | 46.9 | 140 | 48.6 |  |
| Male | 153 | 53.1 | 148 | 51.4 |  |
| **NICU Admission** |  |  |  |  | **33.9 (1), 0.000** |
| NO | 184 | 63.9 | 245 | 85.0 |  |
| YES | 104 | 36.1 | 43 | 15.0 |  |
| **Duration of labour** |  |  |  |  | **6.8 (1), 0.009** |
| <12H | 245 | 85.1 | 265 | 92.0 |  |
| >12H | 43 | 14.9 | 23 | 8.0 |  |
| **PPH** |  |  |  |  | 0 (1), 0.999* |
| Present | 1 | 0.03 | 1 | 0.03 |  |
| Absent | 287 | 99.7 | 287 | 99.7 |  |

**Note:** \* Chi-square values included Yates' continuity correction.

Lastly, investigators made efforts to account for factors associated with maternal Albumin-creatinine ratio among women with preeclampsia (compared to women in the control group) using a multivariable binary logistic regression analysis. Specifically, we fitted a multivariable binary logistic regression model with maternal conditions (1= preeclampsia vs. 2 – pregnant women with no evidence of preeclampsia) as the dependent/outcome variable of interest against a number of selected maternal and immediate neonatal variables. For each variable that was tested against maternal condition of preeclampsia, residual analysis to the outcome variable were made in order to assess for their satisfaction to assumptions of linear model. Specifically, residuals were assessed for *normality, homoscedasticity, linearity, independence*, significant *multi-collinearity* as well as possible *autocorrelation*. Candidate variables for multivariable analysis were considered only if they attained a p-value ≤ 0.2 during univariate analysis. Besides, we also assessed for linear model fitness if effect modifications (statistical interactions) were to be included versus if not. The deciding factor for fitting interaction terms were made only if the variables of interest displayed significant correlation during correlation analysis stage. However, none of the interaction terms gave evidence of statistical significance. All tested interaction terms were therefore discarded in the final candidate model. During initial data exploratory analysis, some variables – (e.g. maternal calendar age, duration of labour) had evidence of significant correlation and even satisfied *linearity* with the outcome variable but failed to attain homoscedasticity. They were thus excluded in the linear model analysis. Table 3 below displays summary statistics of the clinical factors associated with maternal albumin-creatinine ratio among studied women at Amana regional referral hospital in Dar es Salaam, Tanzania.

**Table 3:** Multivariable binary logistic regression analysis of factors associated with maternal urinary albumin-to-creatinine levels among pregnant women with pre-eclampsia at Amana regional referral hospital – Dar es Salaam.

| **Univariate /** | **Crude** | **Analysis** |  | **Multivariable analysis** |  |  |
| --- | --- | --- | --- | --- | --- | --- |
| **Variable name** | **Unadjusted /crude odds ratio** | **95% C.I.** | **P-value** | **Adjusted Odds ratio** | **95% C.I** | **P-Value** |
| **Constant** | **0.28** | **0.09 – 0.41** | **0.001** | **0.04** | **0.03 – 0.05** | **0.000** |
| Gestational age at baseline | 1.03 | 1.01 – 1.04 | 0.000 | 1.00 | 1.00 – 1.00 | 0.99 |
| BMI (kgm<sup>-2</sup>) | 0.72 | 0.68 – 0.77 | 0.000 | 0.968 | 0.69 – 1.71 | 0.881 |
| Apgar score at 1<sup>st</sup> minute | 1.1 | 0.99 – 1.2 | 0.08 | 0.88 | 0.00 – 1.32 | 0.461 |
| **Apgar score at 5<sup>th</sup> minute** | **1.11** | **1.01 – 1.21** | **0.023** | **1.07** | **1.00 – 1.33** | **0.000** |
| **Neonatal birth weight** | **2.15** | **1.6 – 2.9** | **0.000** | **2.0** | **1.2 – 2.9** | **0.000** |
| **GA at delivery** | **1.75** | **1.53 – 2.00** | **0.000** | **1.1** | **1.00 – 1.62** | **0.000** |
| Neonatal admission status | 0.32 | 0.21 – 0.48 | 0.000 | 0.98 | 0.66 – 1.43 | 0.698 |
| **Systolic BP** | **0.26** | **0.13 – 0.53** | **0.000** | **1.01** | **1.00 – 1.77** | **0.009** |
| **Diastolic BP** | **0.66** | **0.61 – 0.71** | **0.000** | **1.12** | **1.00 – 1.97** | **0.003** |
| Gravidity | 1.04 | 0.96 – 1.13 | 0.365 |  |  |  |
| Parity | 1.05 | 0.94 – 1.18 | 0.418 |  |  |  |
| Number of living children | 1.09 | 0.97 – 1.23 | 0.145 | 1.03 | 0.23 – 3.11 | 0.967 |
| **Mode of delivery – Caesarean section** | **0.287** | **0.2 – 0.41** | **0.000** | **1.19** | **1.02 – 1.45** | **0.016** |
| **SVD** | **1** |  |  | **1** |  |  |

## Discussion

We found out that whereas systolic and diastolic blood pressure were significant risk factors associated with *significant proteinuria* among women with preeclampsia compared to those without preeclampsia, Caesarean mode of delivery, 5^th^ minute Apgar score, gestational age at delivery as well as neonatal birth weight to be significant immediate outcomes associated with significant proteinuria among women with preeclampsia compared to those without it, all other factors in check. Specifically, it was interesting to realise that in actual fact, diastolic blood pressure provided a relatively higher significant risk estimate (an increase of about 16%) to systolic blood pressure (an increase of about 5%) towards *significant proteinuria* among studied women with preeclampsia. To the best of our knowledge, the exact reason for such an observation remains elusive but speculated to be a function of renal response to glomerular changes and increased permeability to proteins. Spargo and colleagues coined it as glomerular capillary endotheliosis for the first time in 1976 (22). In fact, it has been shown that glomerular capillary endotheliosis and associated podocyte loss intensity, when severe to result into a nephrotic-range proteinuria (3).

Moreover, we also observed an average increased risk of about 14% for *significant proteinuria* to be observed for each increased week in gestational age at delivery. This finding is consistent with others published before (3, 23). For instance, Wright and colleagues reported their Fetal Medicine Foundation (FMF) competing risk model positive detection rate of around 10% for gestational age at delivery (23). The model’s description towards risk of proteinuria and preeclampsia to gestational age at birth refers to a constellation of attributes (e.g. angiogenic markers, Ultrasonographic assessment of the uterine-artery pulsatility index, maternal BMI as well as maternal racial and ethnic background) (3). Otherwise, the competing risks model of the Fetal Medicine Foundation (FMF) to be supported by the largest body of evidence (3). Furthermore, its statistical vigour is based on a survival-time model that incorporates a prior distribution of gestational age at delivery with preeclampsia, derived from maternal characteristics, with likelihood functions from biomarkers to estimate an individual woman’s associated hazard of delivery with preeclampsia before a specified gestational age (3).

We also derived an estimated increased risk of 24% for *significant proteinuria* to be present among women with preeclampsia who prospectively delivered by Caesarean section. We considered that observation to be a known sequela of fetal manifestations of preeclampsia. Magee and colleagues in their recent New England Journal of Medicine publication reported non-uniform fetal manifestations associated with preeclampsia (both proteinuric and non-proteinuric) to include macrosomia and fetal growth restrictions (3). They evidently narrated whereas macrosomia to be a sequela of utero-placental mismatch, especially in late-onset preeclampsia, fetal growth restriction is a function of inadequate placentation, usually observed in early-onset preeclampsia (3). Other investigators also reported similar risks findings like ours (24).

On neonatal immediate outcomes, we found out an average increased risk of around 11% for significant proteinuria among newborns of preeclamptic women for each unit change in 5^th^ minute Apgar score. Previous studies also reported similar findings (25, 26). that 5^th^ minute Apgar score to be a better predictor (than 1^st^ minute Apgar score) of adverse outcomes in this case, since the index reflects a sustained adaptation (and possible recovery?) of newborns to otherwise adverse in-utero conditions (25, 26).

Our study is the first in Africa to depict the prospective analysis of factors associated with significant proteinuria among women with preeclampsia. Besides, we employed a multivariable linear model and hence the final analysis controlled for both known and even unknown confounders in its estimates. Moreover, the fact that we exclusively applied albumin-to-creatinine ratio further increased chances for characterizing significant proteinuria in preeclampsia. Previous studies have reported albumin to be a better predictor of proteinuria to other serum protein/total protein levels during pregnancy (27). We also acknowledge this study to be part of a locally derived initiatives to the growing demands for local data in clinical research for global consumption as reported before (28). Furthermore, the fact that there are multiple newborn’s based factors associated with maternal urinary albumin-to-creatinine ratio among preeclamptic, suggest possible antecedent cues for the current reported deleterious health consequences among newborns (29, 30) and under-five year population in Tanzania (31-35).

However, our study has several limitations. First, we were limited by design to pre-partum preeclampsia. Moreover, our analysis was derived using linear model. Thus, for all those factors that interact with maternal albumin-to-creatinine ratio in a non-linear fashion was likely penalized. We do accept all setbacks associated with appropriate usage of our fitted model, pending a better model than ours in the future. Our token of advice, with the growing demand for reliable and validated models in clinical medicine (3, 36, 37), we do call for pioneering move in order to achieve personalized medical care in future.

## Data Availability

All data produced in the present study are available upon reasonable request to the authors

## References

1. Khan K, Wojdyla D, Say L, Gulmezoglu A. and van Look P, WHO analysis of the global causes of maternal death: a systematic review. Lancet 2006; 367: 1066–1074.

2. GBD 2015. Maternal mortality collaborators. Global, regional and national levels of maternal mortality, 1990-2015: a systematic analysis for the Global Burden of Disease Study 2015. Lancet 2016; 388: 1775–1812.

3. Magee L, Nicolaides K. and von Dadelszen P. Preeclampsia. N Engl J Med. 2022; 386: 1817–1832.

4. Lindheimer M, Taler S. and Cunningham F. ASH position article: hypertension in pregnancy. J Am Soc Hypertens 2008; 2: 484–489.

5. Tuffnell D, Shennan A, Waugh J. and Walker J. The management of severe preeclampsia/eclampsia, guideline no. 10(A). London: Royal College of Obstetricians/Gynaecologists, 2006.

6. Newman G, Robichaux A, Stedman C, Jaeckle R, Fontenot M, Dotson T, et al. Perinantal outcomes in preeclampsia that is complicated by massive proteinuria. Am J Obstet Gynecol. 2003; 188(1): 264–268.

7. Chan P, Brown M, Simpson J. and Davis G. Proteinuria in preeclampsia: how much matters? BJOG 2005; 112(3): 280–285.

8. Dwyer P, Gorman M, Carroll I. and Druzin M. Urinalysis vs. urine protein-creatinine ratio to predict significant proteinuria. J Perinatol. 2008; 28(7): 461–467.

9. Masini G, Foo L, Tay J, et al. Preeclampsia has 2 phenotypes that require different treatment strategies. Am J Obst Gynecol. 2021;

10. Burton G, Redman C, Roberts J. and Moffett A. Pre-eclampsia: pathophysiology and clinical implications. BMJ 2019; 366: 2381

11. Lamminpaa R, Vehvilainen-Julkunen K, Gissler M. and Heinonen S. Preeclampsia complicated by advanced maternal age: a registry-based study of primiparous women in Finland 1997-2008. BMC Pregnancy & Childbirth 2012: 12: 47.

12. Kuo VS, Koumantakis G, Gallery EDM. Proteinuria and its assessment in normal and hypertensive pregnancy. Am J Obstet Gynecol. 1992;167(3):723–8.

13. Naruse M, Mukoyama M, Morinaga J, Miyazaki M, Iseki K, Yamagata K. Usefulness of the quantitative measurement of urine protein at a community-based health checkup: a cross-sectional study. Clin Exp Nephrol. 2020;24(1):45–52.

14. Ransohoff D. and Feinstein A. Problems of spectrum and bias in evaluating the efficacy of diagnostic tests. N Engl J Med. 1978; 299: 926–930.

15. Knottnerus J. and Muris J. Assessment of the accuracy of diagnostic tests: the cross-sectional study. J Clin Epidemiol. 2003; 56: 1118–1128.

16. Lijmer J, Mol B, Heisterkamp S, Bonsel G, Prins M, van der Meulen J, et al. Empirical evidence of design-related bias in studies of diagnostic tests. JAMA 1999; 282(11): 1061–1066.

17. Durnwald C. and Mercer B. A prospective comparison of total protein/creatinine ratio versus 24-hour urine protein in women with suspected preeclampsia. Am J Obstet Gynecol. 2003; 189: 848–852.

18. Leshabari K. Demographic transition in sub-Saharan Africa: from grassroots to ivory towers. In Klimczuk (ed) Demographic analysis – selected concepts, tools and applications. InTech Open: London, 2021. Available from [Accessed on 1 October 2025]

19. Leshabari K, Biswas A, Gebuis E, Leshabari S. and Ohnishi M. Challenges in morbidity and mortality statistics of the elderly population in Tanzania: a call to action. Quality in Ageing and Older Adults 2017; 18(3): 171 – 174.

20. Machano M. and Joho A. Prevalence and risk factors associated with severe pre-eclampsia among postpartum women in Zanzibar: A cross-sectional study. BMC Public Health 2020;20(1):1–10.

21. Mboya T, Chiduo M, Sylvester B. and Leshabari K. Diagnostic test performance of urinary albumin-to-creatinine ratio for detection of significant proteinuria in preeclampsia. BMJ 2026;

22. Spargo B, Lichtig C, Luger A, Katz A. and Lindheimer M. The renal lesion in preeclampsia. Perspect Nephrol Hypertens. 1976; 5: 129–137.

23. Wright D, Wright A, Nicolaides K. The competing risk approach of preeclampsia. Am J Obst Gynecol. 2020; 223(1): 12–23.

24. Lei T, Qiu T, Liao W, Li K, Lai X, Huang H, et al. Proteinuria may be an indicator of adverse pregnancy outcomes in patients with preeclampsia. Reproductive Biology and Endocrinology 2021; 19:71.

25. Cnattingius S, Norman M, Granath F, Peterson G, Stephansson O. and Frisell T. Apgar score components at 5 minutes: Risks and prediction of neonatal mortality. Pediatric and Perinatal Epidemiology 2017; 31(4): 328–337.

26. Ehrhardt H, Behboodi S, Maier R, Aubert A, Aden U, Draper E, et al. Five-minute Apgar scores and its prognostic value for mortality and severe morbidity in very preterm infants: A multinational cohort study. BJOG 2025;

27. Bartal M, Lindheimer M, Sibai B. Proteinuria during pregnancy: definition, pathophysiology, methodology, and clinical significance. Am J Obst Gynecol. 2022; 23: S819–S834.

28. Swai G. and Leshabari K. The new paradigm of local research and innovations by local health professionals. Tanzania Journal of Health Research 2022; 23. Available from https://scholar.google.com/citations?view_op=view_citation&hl=en&user=6GgGNakAAAAJ&citation_for_view=6GgGNakAAAAJ:WqliGbK-hY8C [Accessed on 11 November 2025]

29. Mhando G, Kalabamu S, Fataki M, Galabawa C. and Leshabari K. Predictors of early newborn deaths at Dar es Salaam public regional referral hospitals: A prospective observational hospital-based study. PLoS ONE 2025; 20(12): e0338497. Available from https://journals.plos.org/plosone/article?id=10.1371/journal.pone.0338497 [Accessed on 26 December 2025]

30. Salum S, Kalabamu F, Fataki M, Omary S, Mohammed U, Kizwi H, et al. Asymptomatic hypoglycemia among preterm newborns: a cross-sectional analysis. PLoS ONE 2024; 19(4): e0301803. Available from https://journals.plos.org/plosone/article?id=10.1371/journal.pone.0301803 [Accessed on 10 October 2025]

31. Leshabari K. and Ramji R. The pattern of infections among under-fives: a call for actions. Dares Salaam Medical Students’ Journal 2008; 15(1): 24–29.

32. Omary S, Kalabamu F, Fataki M, Salum S, Mohamed U, Kimaro J, et al. Severity and morphological characteristics of anaemia among 6 to 59 months old children in Temeke, Dar es Salaam – Tanzania: clinics-based cross-sectional analysis. East African Health Research Journal 2024; 8(2): 188–194.

33. Jumanne S, Meda J, Hokororo A, Leshabari K. Clinical predictors of malaria, acute bacterial meningitis and treatment outcomes among febrile children admitted with altered mental status in Northwestern Tanzania. J Trop Pediatr. 2018; 64(5): 426–433.

34. Salum S, Kalabamu F, Fataki M, Omary S, Mohamed U, Kizwi H, et al. Prevalence and factors associated with asymptomatic hypoglycemia among preterm newborns in Dar es Salaam regional referral hospitals, Tanzania: a cross-sectional analytical study protocol. MedRxiv. Available from https://www.medrxiv.org/content/10.1101/2022.10.28.22281650v1 [Accessed on 26 December 2025]

35. Omary S, Kalabamu F, Fataki M, Salum S, Mohamed U, Kimaro J, et al. Severity and morphological classification of anaemia among children aged 2-59 months in Dar es Salaam, Tanzania: a cross-sectional study protocol. MedRxIV. Available from https://www.medrxiv.org/content/10.1101/2022.11.10.22282169v1 [Accessed on 26 December 2025]

36. Leshabari K. Reliability and validity of clinicopathological features associated with frailty syndrome in elderly population. In Palermo S (ed) Frailty in the elderly Understanding and managing complexity. IntechOpen: London, 2021. Available from https://www.intechopen.com/chapters/73143 [Accessed on 10 October 2025]

37. Leshabari K, Ndonde W, Mbega S, Mbululo L. Predictors of cardiometabolic risks among typical African elderly population: Analysis from Dar es Salaam, Tanzania. Diabetes Technology & Therapeutics 2020; 22: A214.

